# Pregnancy outcomes and HIV amongst women in Northwest Tanzania, from 2015-2016

**DOI:** 10.1101/2025.01.31.25321464

**Authors:** Samika Hariharan, Neema Mosha, Jacqueline Materu, Margaret Baker, Coleman Kishawame, Mark Urassa

## Abstract

**Background:** Adverse pregnancy outcomes are rampant across Sub Saharan Africa and significantly increase the risk of neonatal deaths. HIV infection is a key contributor to adverse pregnancy outcomes and has a disproportionately high burden amongst women in both Sub Saharan Africa and Tanzania. Socio-demographic factors and care-seeking behaviors can either exacerbate or prevent HIV infection and adverse pregnancy outcomes.

**Objectives:** This study aims to determine HIV prevalence by age, residence, marital status, and education and to determine how pregnancy outcomes are impacted by HIV infection, the use of health services, and socio-demographic factors.

**Methods:** This study utilized secondary data from an epidemiological Sero Survey Round 8 (2015-2016) within the Kisesa HIV cohort in Magu District, Mwanza, Tanzania. All data was analyzed using the statistical software “Stata Version 17.0.” The variables were re-coded for operational use and cross-tabulations with chi-squared tests and logistic regression models were conducted.

**Results:** Data analysis showed an HIV prevalence of 9.5% amongst women who had been pregnant before. HIV positive women were more likely to lose a child (AOR=1.4; 95% CI: 1.1,1.7). Women with higher education had significantly lower odds of adverse pregnancy outcomes as compared to women with no formal education (AOR=0.5; 95% CI: 0.4, 0.7). Women with more children, especially 10+ children, also had higher odds of adverse outcomes as compared to mothers with 1-3 children (AOR=25.1; 95% CI: 17.0, 37.0). Elderly mothers had seven times the odds of losing a child as compared to young mothers (AOR=7.0; 95% CI: 2.5, 19.6).

**Conclusions:** This study provides needed insight into factors that contribute to HIV infection and adverse pregnancy outcomes. The results support the implementation of new policies that target HIV prevention and treatment amongst women and emphasize maternal education to reduce adverse pregnancy outcomes.

## Background

The improvement of maternal, reproductive, and child health in Tanzania has been a major goal for the country since the early 21^st^ century. Neonatal mortality contributes to 40% of under-five (U5) child mortality in Tanzania [1]. Preterm deliveries have been shown to contribute to one quarter of neonatal deaths across the world [2]. Tanzania has over 300,000 preterm deliveries every year and is among the top ten countries with the highest number of preterm births [2]. Various factors have been shown to decrease preterm births and other adverse pregnancy outcomes, including higher-level education of women, higher socioeconomic status, and more antenatal care follow-ups [3]. Conversely, intimate-partner violence, multiparity, anemia during pregnancy, and a lack of involvement of women in the healthcare decision making process have been associated with increased adverse outcomes [2, 4].

HIV infection has also been shown to significantly exacerbate both neonatal and maternal mortality [5]. HIV-positive women who become pregnant have an increased risk of adverse pregnancy outcomes including spontaneous miscarriages, stillbirths, and intrauterine growth restriction of the fetus [5]. HIV positive pregnant women also risk vertical transmission, which is the spread of HIV from an HIV positive mother to her child during pregnancy, labor, or breastfeeding. Without treatment, anywhere between 15-45% of HIV positive women will transmit HIV to their babies [6]. If HIV is transmitted, one third of HIV-positive children in SSA will die before their first birthday, and over one half will die before their second birthday [7]. Given the high burden of HIV on pregnancy outcomes, HIV infection was identified as a key field of study to improve reproductive and child health in Tanzania.

Across the globe, 38.4 million people are living with HIV (PLHIV) and in 2022, 630,000 HIV-infected people died due to AIDS-related illnesses [8]. However, the burden of HIV is not evenly distributed. While only 0.5% of people in North America have HIV, 67% of the world’s population of PLHIV are in Sub Saharan Africa (SSA) [9–10]. Women and girls face a disproportionately high burden of HIV in SSA and 95% of HIV-positive women live in SSA [7]. HIV prevalence is higher amongst women because of a host of factors; women have thin vaginal mucus that increases the likelihood of HIV transmission, patriarchal norms and gender-based violence allow men to dictate when and how women will have sex, and many young women resort to transactional sex for cash transfers [11].

Tanzania is one of the countries in SSA most impacted by the HIV/AIDS epidemic. Within countries in Africa, Tanzania has the 5^th^ highest prevalence of HIV, with an HIV prevalence of approximately 4.7% [12–13]. HIV prevalence is higher amongst women (6.2%) than men (3.7%) [14]. Unfortunately, 25% of HIV positive women in Tanzania will still transmit HIV to their babies [14]. HIV positive women in Tanzania have also been shown to have lower overall fertility (4.4) as compared to HIV negative women (6.7) [15].

ART, or antiretroviral therapy, is an HIV treatment regimen that decreases the lethal effects of HIV and makes PLHIV non-infectious [16–18]. Prevention of mother-to-child transmission (PMTCT) is a form of ART for pregnant women that can decrease vertical transmission rates to below 1% [7]. In SSA, nearly 50% of HIV-positive pregnant women fail to access PMTCT [7]. Low PMTCT utilization is often due to poor knowledge about vertical transmission and PMTCT, inadequate access to PMTCT, and heavy HIV stigma [7].To combat the low utilization of ART and PMTCT, HIV Counseling and Testing (HCT) has been shown to be effective in educating people on HIV, HIV transmission mechanisms, and one’s HIV status [19]. In turn, this knowledge has been shown to decrease risky sexual behaviors [20]. HCT also helps to guide PLHIV through the lifesaving ART process [19].

In Tanzania, efforts to integrate PMTCT into maternal, neonatal, and child health have allowed for approximately three quarters of women with HIV to receive PMTCT and prevent vertical transmission [21]. This process has required more decentralization of PMTCT services from larger hospitals to local primary care centers [22]. Despite decentralization, there are many structural factors that decrease the utilization of critical services by the 70% of Tanzanians that live in rural areas [23–24]. These factors include a shortage of antenatal care clinics, longer and more expensive travel, lower socioeconomic status, less than 40% of the required health workforce, and a severe lack of comprehensive obstetric care [23–25]. In turn, less utilization of services––including 30% less utilization of birthing in skilled facilities––can worsen pregnancy outcomes for women in rural areas [24].

Given the seriousness of the burden of HIV infection on women in SSA, as well as the burden that HIV infection has on pregnancy outcomes, this study focused on both HIV and pregnancy outcomes in Tanzania. This research is important because it addresses gaps in research HIV and pregnancy outcomes, specifically providing insight into the situation in Tanzania and the Mwanza region. This study also conducts a uniquely comprehensive analysis of variables that impact pregnancy outcomes in Mwanza, exploring factors from sociodemographic variables to care-seeking behaviors to HIV status.

This paper aims first to understand the factors associated with HIV prevalence and second to understand factors associated with pregnancy outcomes among women who have ever been pregnant in the Mwanza region of Northwest Tanzania. This information is critical for the design of interventions to reduce HIV among women and to improve pregnancy outcomes.

## Methodology

### Data collection

The data for this study was obtained from an epidemiological sero-survey implemented during 2015-2016 within the Magu Health and Demographic Surveillance System (HDSS). The Magu HDSS is also known as the Kisesa observational HIV cohort in Mwanza Region, Northwest Tanzania. Details regarding Magu HDSS survey methodology were published in the International Journal of Epidemiology [26].

### Inclusion criteria

Figure 1 shows how the total study population (n=10,916 participants) was narrowed down to the target sample (n=4,096) by excluding men (n=4,084), women who had never been pregnant (n=1,456) and participants who did not answer the relevant survey questions (n=1,820).

**Figure 1.**
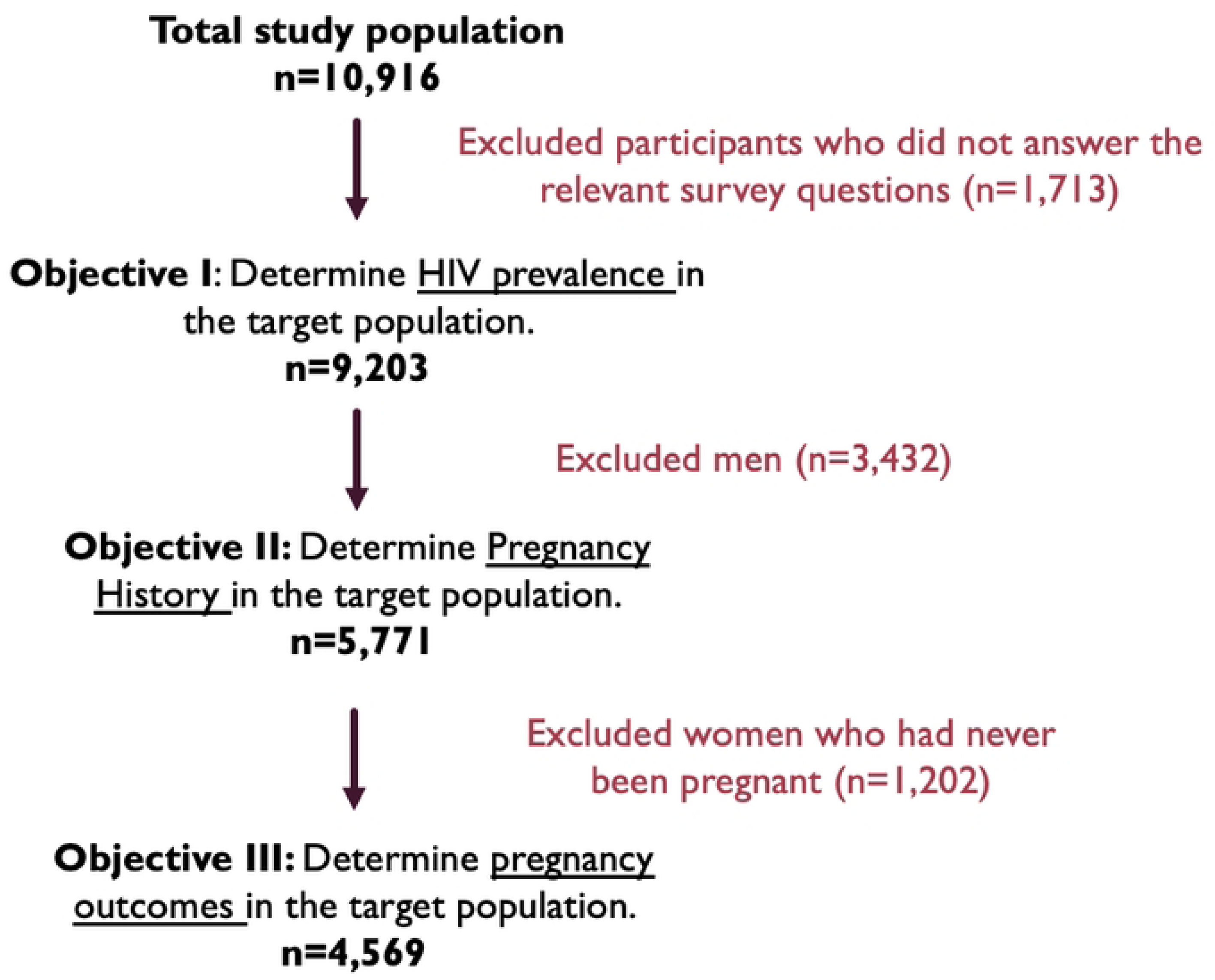

### Objectives

The study objectives were two-fold: Firstly, to determine HIV prevalence by age, education level, marital status, and residence status amongst women who have been pregnant in Northwest Tanzania. Second, to determine how pregnancy outcomes are influenced by HIV Status, HIV care-seeking behaviors, and demographic characteristics (residence, age, education level, and number of children). In all, the study sought to answer the question: What are key sociodemographic and care-seeking factors that can be used to improve the burden of HIV and pregnancy outcomes amongst women in Northwest Tanzania?

### Study variables

Relevant demographic questions covered age, village of residence, sex, marital status, occupation, education level, and number of children. Other relevant survey questions asked about pregnancy history

(losses, still-births, miscarriages), use of HCT services, and health service use in the past year. HIV status was determined from lab results. **Table 1** shows the recoding and categorization of study variables [27–29].

**Table 1:** Recording and Categorization of the Variables

| Variable | Description | Categories |
| --- | --- | --- |
| Pregnancy Outcomes | Number of miscarriages, still births, or loss of children after birth that the participant has experienced | No Children Lost<br>Spontaneous Miscarriages<br>Still Births<br>1+ Children Lost After Live Birth |
| Number of Children | Participant's total number of live births as categorized by collapsing United Nations categories [27] | 1 - 3, 4 - 5, 6 - 9, or 10+ Children |
| HIV Status | Participant's HIV status as determined by lab results | HIV Negative<br>HIV Positive |
| HCT Use | Has the participant used HIV Counseling and Testing (HCT) before? | Yes or No |
| General Health Service Use | Participant's frequency of use of health services in the last year | "Infrequent Use" for 0-3 visits in the last year<br>"Moderate Use" for 4-7 visits in the last year<br>"Consistent Use" for 8+ visits in the last year |
| Residence | Participant's residence area | Rural [Igekemaja or Kitumba or Isangijo or Ihayabuyaga or Welamasonga] or Semi-urban [Kanyama or Kisesa or Wita or Bujora] |
| Education | Level of education that the participant has either started or completed | No formal education<br>Attended or Completed Primary School<br>Attended or Completed Secondary School or Higher Education |
| Age | Participant's age group classification per the National Institute of Health's Age Group Classifications [28] | Young Women (15-29)<br>Middle-Aged Women (30-44)<br>Older Women (45-59)<br>Elderly Women (60+) |
| Marital Status | Marital status of participant | Never Married<br>Monogamously Married<br>Polygamously Married<br>Widowed<br>Separated / Divorced |
| Occupation | Participant's occupation per the International Standard Classification of Occupations [29] | Students, Individuals Who Do Not Work, Individuals who Perform Unskilled Labor Jobs, Individuals who Performed Skilled Work, and Self-Employed Individuals (including business owners and professionals) |

### Data analysis

All data was analyzed using “Stata Version 17.0” (Stata Corporation, College Station, TX, USA). After recoding, the frequencies of each variable were tabulated. Bivariate analysis was performed using Chi-square tests to determine the prevalence of HIV by sociodemographic characteristic of residence, education, age, and marital status. Similar analysis was done but with pregnancy outcomes as the outcome variable. Pregnancy outcomes were analyzed as a function of education level, age, residence, number of children, HIV status, and care-seeking behaviors. Logistic regression models were used to determine the association between pregnancy outcomes and the defined exposure variables. Prior to the logistic regression analysis, the primary outcome variable of pregnancy outcomes was collapsed into two categories (“No Losses” and “Lost 1 or More Children”). All analyses were considered significant at a P-value < 0.05.

### Ethical considerations

The original survey study obtained informed consent from all its participants and provided follow-up testing, counseling, and treatment to better participants’ health following the survey. The Sero-Survey received approval from the Tanzania Medical Research Review Committee, through a certificate with reference number: NIMR/HQ/R.8a/Vol.IX/1489 The current study conducted secondary data analysis using anonymized data from the Sero-Survey Round 8. No contact was made with the original survey participants. As such, no consent needed to be obtained.

Data was accessed from 27 October 2023 to 26 October 2024. A confidentiality contract was signed by the researchers to ensure that the data was stored safely and privately and removed from personal devices following study completion. The study received ethical clearance from the Tanzania Medical Research Coordinating Committee, with certificate number: NIMR/HQ/R.8a/Vol.IX/4444.

## Results

**Table 2** shows socio-demographic characteristics of target study participants. There were 4,096 females studied. 33.5% had not received any formal education while only 10% had completed secondary or higher education. In terms of occupation, nearly one quarter of the women (23.8%) did not work, while 70.2% of women were involved in some form of unskilled labor. 61.2% of the population was in monogamous marriages and 7.8% was polygamously married. Participants were primarily aged 15-29 years old (38.7%) and 30-44 years old (35.4%). Less than 10% (8.3%) were over 60 years old. The median age of the survey participants was 33 years.

**Table 2:** Socio-Demographic Characteristics of the Target Sample of Female Participants who have been Pregnant Before (N=4,096)

|  | **N= 4,096**<br>**n (%)** |  |
| --- | --- | --- |
| **Residence Status** |  |  |
| Semi-Urban | 2,066 | (50.4) |
| Rural | 2,030 | (49.6) |
| **Education** |  |  |
| None | 1,373 | (33.5) |
| Primary | 2,314 | (56.5) |
| Secondary or Higher | 409 | (10.0) |
| **Employment Status** |  |  |
| Student | 36 | (0.9) |
| Doesn't Work | 976 | (23.8) |
| Unskilled Labour | 2,877 | (70.2) |
| Skilled Labour | 1 | (0.0) |
| Self-Employed | 206 | (5.0) |
| **Marital Status** |  |  |
| Never Married | 312 | (7.6) |
| Monogamously Married | 2,507 | (61.2) |
| Polygamously Married | 321 | (7.8) |
| Widowed | 412 | (10.0) |
| Separated / Divorced | 544 | (13.3) |
| **Age (Years)** |  |  |
| Young Women (15-29) | 1,584 | (38.7) |
| Middle-Aged Women (30-44) | 1,452 | (35.4) |
| Older Women (45-59) | 721 | (17.6) |
| Elderly Women (60+) | 339 | (8.3) |
|  | **Median Age (IQR)** |  |
|  | 33 (15 – 91) |  |

**Table 3** presents HIV status by socio-demographic characteristics. Nearly ten percent (n=398, 9.5%) of the women studied were HIV positive. Differences in HIV prevalence by residence was not statistically significant (P=0.142). HIV prevalence was highest among those with no formal education (12.2%) and lowest amongst those with higher education (3.7%) (P <0.001). There was also a significant difference in HIV prevalence by the age of participants; HIV prevalence was 13.4% among participants in the middle-age group and 11.0% in the elderly age group, and there were lower prevalences of HIV in the youngest (6.1%) and oldest (5.6%) age groups (P <0.001). Separated or divorced women had the highest HIV prevalence at 15.4%, and women who were monogamously married had the lowest HIV prevalence at 7.1% (P<0.001).

**Table 3:** HIV Status by Socio-Demographic Characteristics (N=4,096)

|  | HIV Negative | HIV Positive | Total | P-Value* |
| --- | --- | --- | --- | --- |
|  | N= 3,707 (90.5%) | N= 389 (9.5%) |  |  |
|  | n (%) | n (%) | n |  |
| **Residence Status** |  |  |  |  |
| Semi-Urban | 1,856 (89.8) | 210 (10.2) | 2,066 | 0.142 |
| Rural | 1,851 (91.2) | 179 (8.8) | 2,050 |  |
| **Education** |  |  |  |  |
| None | 1,205 (87.8) | 168 (12.2) | 1,373 | < 0.001 |
| Primary | 2,108 (91.1) | 206 (8.9) | 2,314 |  |
| Secondary or Higher | 394 (96.3) | 15 (3.7) | 409 |  |
| **Age (Years)** |  |  |  |  |
| Young Women (15-29) | 1,488 (93.9) | 96 (6.1) | 1,584 | < 0.001 |
| Middle-Aged Women (30-44) | 1,257 (86.6) | 195 (13.4) | 1,452 |  |
| Older Women (45-59) | 642 (89.0) | 79 (11.0) | 721 |  |
| Elderly Women (60+) | 320 (94.4) | 19 (5.6) | 339 |  |
| **Marital Status** |  |  |  |  |
| Never Married | 277 (88.78) | 35 (11.22) | 312 | < 0.001 |
| Monogamously Married | 2,329 (92.90) | 178 (7.10) | 2,507 |  |
| Polygamously Married | 291 (90.65) | 30 (9.35) | 321 |  |
| Widowed | 350 (84.95) | 62 (15.05) | 412 |  |
| Separated / Divorced | 460 (84.56) | 84 (15.44) | 544 |  |

\*P-value measures if there is a significant difference in HIV prevalence as a function of sociodemographic characteristics.

**Table 4** shows pregnancy outcomes in relation to demographic variables, HIV status, and care-seeking behaviors. As education level increased, stillbirths and losses after birth decreased. The proportion of women with no losses similarly increased with education level, from 43.9% to 79.5%. Higher maternal age during pregnancy also increased with adverse outcomes. For example, more elderly mothers lost children after their last pregnancy (21.4%) as compared to 2.8% of young mothers. Finally, as a mother’s number of children increased, the percentage of mothers experiencing no losses decreased from 77.5% for mothers with 1-3 children to 9.2% for those with 10+ children. All demographic variables were significantly associated with pregnancy outcomes (P<0.001).

**Table 4:** Pregnancy Outcomes by HIV Status, Demographic Variables, and Care-Seeking Behaviors (N=4,096)

|  | Pregnancy Outcome(s) |  |  |  | Total<br>n (%) | P-Value |
| --- | --- | --- | --- | --- | --- | --- |
|  | No Losses<br>n (%) | Stillbirth(s)<br>n (%) | Miscarriage(s)<br>n (%) | Lost<br>Child(ren)<br>After Birth<br>n (%) |  |  |
| Demographic Variables |  |  |  |  |  |  |
| Residence Status |  |  |  |  |  |  |
| Semi-Urban | 1,174 (56.8) | 142 (6.9) | 299 (14.5) | 251 (21.8) | 2,066 | <0.001 |
| Rural | 1,156 (57.0) | 65 (3.2) | 244 (12.0) | 565 (27.8) | 2,030 |  |
| Education |  |  |  |  |  |  |
| None | 603 (43.9) | 86 (6.3) | 185 (13.5) | 499 (36.3) | 1,373 | <0.001 |
| Primary | 1,402 (60.6) | 111 (4.8) | 317 (13.7) | 484 (20.9) | 2,314 |  |
| Secondary or Higher | 325 (79.5) | 10 (2.4) | 41 (10.0) | 33 (8.1) | 409 |  |
| Age at Last Pregnancy (Years) |  |  |  |  |  |  |
| Young Women (15-29) | 3,101 (95.3) | 14 (0.4) | 47 (1.4) | 91 (2.8) | 3,253 | <0.001 |
| Middle-Aged Women (30-44) | 531 (89.4) | 3 (0.5) | 7 (1.2) | 53 (8.9) | 594 |  |
| Older Women (45-59) | 183 (82.8) | 0 (0.0) | 0 (0.0) | 38 (17.2) | 221 |  |
| Elderly Women (60+) | 22 (78.6) | 0 (0.0) | 0 (0.0) | 6 (21.4) | 28 |  |
| Number of Children |  |  |  |  |  |  |
| 1-3 | 1,411 (77.5) | 46 (2.5) | 192 (10.6) | 171 (9.4) | 1,820 | <0.001 |
| 4-5 | 505 (55.7) | 61 (6.7) | 118 (13.0) | 222 (24.5) | 906 |  |
| 6-9 | 382 (37.3) | 65 (6.4) | 147 (14.4) | 429 (41.9) | 1,023 |  |
| 10+ | 32 (9.2) | 35 (10.1) | 86 (15.8) | 194 (55.9) | 347 |  |
| HIV Status |  |  |  |  |  |  |
| Negative | 2,138 (57.7) | 186 (5.0) | 486 (13.1) | 897 (24.2) | 3,707 | 0.013 |
| Positive | 192 (49.4) | 21 (5.4) | 57 (14.7) | 119 (30.6) | 389 |  |
| Care-Seeking Behaviours |  |  |  |  |  |  |
| HCT Use |  |  |  |  |  |  |
| No | 731 (56.7) | 70 (5.4) | 141 (10.9) | 348 (26.7) | 1,290 | 0.008 |
| Yes | 1,599 (57.0) | 137 (4.9) | 402 (14.3) | 668 (23.8) | 2,806 |  |
| General Health Service Use |  |  |  |  |  |  |
| Infrequent | 992 (56.7) | 83 (4.8) | 205 (11.7) | 469 (26.8) | 1,749 | 0.012 |
| Moderate | 566 (56.3) | 45 (4.5) | 141 (14.0) | 254 (25.3) | 1,006 |  |
| Consistent | 772 (57.6) | 79 (5.9) | 197 (14.7) | 293 (21.9) | 1,341 |  |

\*P-value measures if there is a significant difference in pregnancy outcomes as a function of the exposure variables.

HIV status was also significantly associated with pregnancy outcomes (P=0.013), with 57.7% of HIV negative mothers losing no children as compared to 49.4% of HIV positive mothers. Only 24.2% of HIV negative mothers lost children after birth as compared to 30.6% of HIV positive mothers.

Although pregnancy outcomes were also found to be significantly associated with residence area, HCT use, and general health service use, no clear trends were observed between pregnancy outcomes and these exposure variables.

**Table 5** shows univariate and multivariable logistic regression results. Education level, age at last pregnancy, number of children, and HIV status were significantly associated with pregnancy outcomes. Adjusted logistic regression showed that individuals with secondary education or higher, as well as those with primary education, were less likely to lose children compared to those with no education (AOR=0.5; 95% CI: 0.4, 0.7) and (AOR=0.8; 95% CI: 0.7, 0.9) respectively. The results also showed the highest odds of losing children among elderly women as compared to younger women (AOR=7.0; 95% CI: 2.5, 19.6). A higher number of children was similarly associated with adverse outcomes; Women with 10+ children had 25.1 times the odds of losing children as compared to women with three or less children (AOR=25.1; 95% CI: 17.0, 37.0). HIV positive women had 1.4 times the odds of losing children as compared to HIV negative women (AOR=1.4; 95% CI: 1.1,1.7).

**Table 5:**
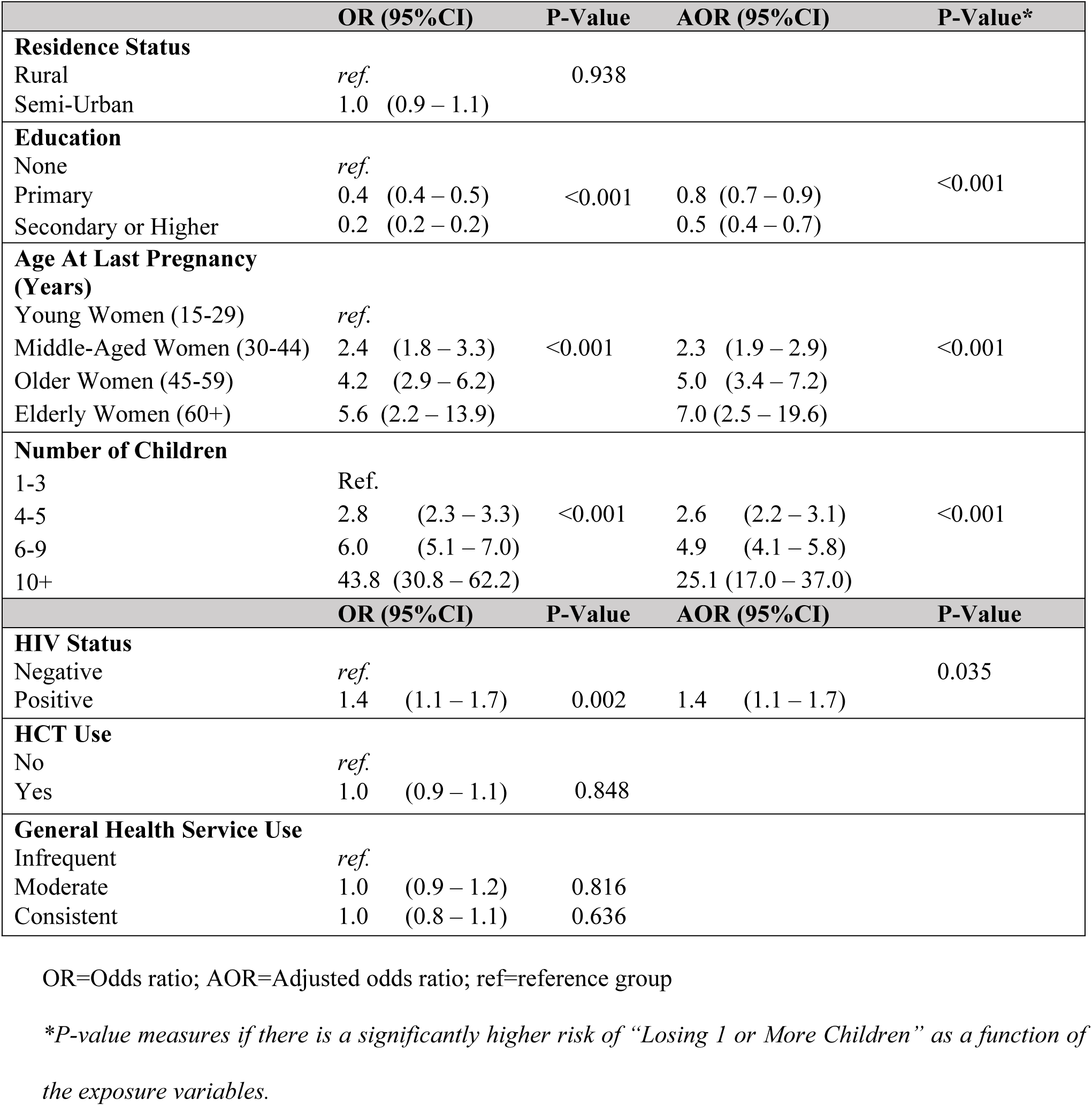
Logistic Regression Associations between Dichotomized Pregnancy Outcomes (“No Losses” vs “Lost 1 or More Children”) and Sociodemographic Variables, HIV Status, and Health Service Use (N=4,096)

## Discussion

The study reported an HIV prevalence of 9.5% amongst women who have been pregnant before. The UNAIDS Tanzania 2020 report showed that there is a higher HIV prevalence of 6.2% amongst women in Tanzania as compared to a prevalence of 4.6% in the general population [30]. Additionally, previous results published about the Magu HDSS have shown that the HIV prevalence amongst the cohort trends similarly to the HIV prevalence from national surveys in Tanzania [26]. Thus, in reporting an HIV prevalence of 9.5%, this study supports that the HIV prevalence amongst women in Tanzania who have been pregnant before is likely higher than the HIV prevalence amongst the general population of women in Tanzania. These results indicate a need for greater HIV prevention and treatment amongst women who have been pregnant before.

The study showed that HIV infection had a negative association with participants’ level of education (P<0.001), with more educated individuals having a lower HIV prevalence. These results are in line with what has been reported from other East African countries [31], but differs from results published in a Tanzanian health survey that reported that those with higher education were 50% more likely to be infected with HIV [32]. More research is needed to understand the relationship between HIV status and education in SSA.

A significant association between age and HIV status (P<0.001) was found in this study. More specifically, middle-aged women aged 30-44 years had the highest HIV prevalence. Other literature reports a similar trend between HIV status and age, with a peak HIV prevalence amongst individuals aged 40-49 years [13]. This spike in HIV amongst middle-aged adults could be impacted by relative practices of polygyny throughout life; Research has shown that polygynous relationships in SSA can increase one’s risk of HIV due to high-risk sexual practices [33]. As such, it is likely non-coincidental that the study’s results showed the highest HIV prevalence amongst participants aged 30-44 years, when Tanzanians aged 35-49 years report the highest rates of polygynous relationships [34]. Study results similarly showed a higher prevalence of HIV in women in polygamous marriages (9.35%) as compared to women in monogamous marriages (7.10%) (P<0.001).

The study also reported the highest HIV prevalence amongst women who had been separated or divorced (15.55%) and widowed women (15.05%). Similar results were reported by a study conducted in South Africa, with widowed women having nearly twice the risk of HIV infection as compared to married women [34]. Thus, the relationship between marital status and HIV prevalence in the Mwanza region is similar to that found elsewhere in SSA.

The demographic variable of education level was found to have a significant relationship with pregnancy outcomes, both through chi squared and logistic regression analysis (P<0.001). Women with higher education had the lowest odds of losing children as compared to women with no formal education (AOR=0.44; 95%CI: 0.34, 0.58). These results align with current literature that shows that educated mothers have more birth preparedness, thereby reducing known risk factors for neonatal death [36–37]. These results provide evidence to support the education of women to improve pregnancy outcomes.

The demographic variable of maternal age during pregnancy was also found to have an association with adverse pregnancy outcomes (P<0.001). Adjusted logistic regression showed that elderly mothers had the highest odds of losing children as compared to young mothers (AOR=25.1; 95% CI: 17.0, 37.0).

Similarly, a longitudinal study in Dar Es Salaam, Tanzania showed that prenatal complications and stillbirths were lowest for teenagers and highest for 35–50-year-old women [38]. These results align with the current study’s results but fail to corroborate medical research that suggests that young mothers are also high-risk for adverse pregnancy outcomes due to their smaller pelvic bone structures that can result in obstructed labor [39–40]. Further research should be conducted in Tanzania to determine if young mothers are at risk for adverse pregnancy outcomes.

HIV status was also significantly associated with adverse pregnancy outcomes among the study participants, with HIV positive women having 1.24 higher odds of losing a child (AOR=1.24; 95% CI: 1.02,1.52) as compared to HIV negative women. A study in Northeast Tanzania published similar results, with HIV positive mothers having over 1.5 times the odds of perinatal death [41]. Studies from Dar Es Salaam and Lesotho also corroborate these results, showing that the relationship between HIV and pregnancy outcomes in Northwest Tanzania is like that across SSA [42–43].

A positive correlation was also found between a woman’s number of children and adverse pregnancy outcomes, with women with 10+ children having 25.1 times the odds of adverse outcomes (AOR=25.1; 95% CI: 17.0, 37.0) as compared to women with 1-3 children. These results are supported by another study conducted in Northern Tanzania that revealed that grand multiparity, defined as a parity of five or more deliveries, was significantly associated with adverse pregnancy outcomes [44].

Although chi squared analysis supported a significant difference in pregnancy outcomes as a function of residence, general health service use, and HCT use, logistic regression that controlled for other variables did not yield significant results for these exposure variables. First looking at residence status, Tanzania is the only country in SSA where neonatal mortality is higher in urban areas [24, 45]. Notably, women in rural Tanzania utilize less health services. Thus, it has been hypothesized that this difference in neonatal mortality could be due to a greater registration of mortality in urban settings [46]. Next, although no significant relationship was found between general health service use and pregnancy outcomes, current literature supports that antenatal care usage should decrease adverse outcomes for mothers [47–48]. Finally, by encouraging PMTCT use, HCT should logically improve pregnancy outcomes by preventing HIV-related adverse events [49]. However, there is little literature beyond this study that directly analyzes the effect of HCT use on pregnancy outcomes. More research should be conducted to determine how the variables studied may affect pregnancy outcomes.

### Strengths and limitations

This study has some limitations worth mentioning: Given that the secondary analysis was based on Sero-Survey Round 8 interviews that asked participants to recall information from their past, the data is subject to both recall bias and response bias. Additionally, the Sero-Survey did not ask questions about the nature of one’s health visits (i.e. preventative or curative). Thus, although preventative health service use could have decreased adverse health outcomes, repeated health service use could also be indicative of severe health issues. This unaccounted-for difference could have contributed to the insignificant results found for the “General Health Service Use” variable during logistic regression. Finally, although this study reported findings amongst a target population of women who had been pregnant before, little other current literature uses a similar reference population. The difference in study populations reduces the ability of this study to be compared to other literature, while also revealing a need for more research amongst the study population of women who have been pregnant before. Finally, residence type designations of rural and semi-urban could not ensure that each category had purely rural or semi-urban elements. This overlap also limits the study’s ability to compare its residence-focused results to other literature.

This study’s strengths lie in its ability to analyze self-reported information from women about their pregnancy histories and child survival, of which there is limited literature. Key findings can guide both future research and policy to improve pregnancy outcomes and decrease the burden of HIV in Tanzania.

## Conclusion and future directions

This study adds to the growing body of research on HIV infection, providing needed insight into the relationship between pregnancy outcomes, HIV infection, and socio-demographic variables in Northwest Tanzania. Firstly, the study offers further evidence to suggest that the prioritization of HIV care for women would not only significantly decrease the burden of HIV in Tanzania but also improve pregnancy outcomes. Second, future studies should explore how interventions can reduce practices of polygamy amongst middle aged individuals, as well as sexual practices amongst widowed or divorced individuals. Finally, the study provides evidence to support that the general and health-specific education of mothers can minimize adverse pregnancy outcomes––especially those that arise from the poor childbearing practices of elderly mothers and mothers of grand multiparity.

In the future, large, longitudinal, prospective, and experimental studies should be conducted to allow for real-time and updated analysis of the variables studied. Such studies may inform future policies around maternal health services and HIV in Tanzania.

## Data Availability

The Magu HDSS data that was analyzed can be found at the National Institute for Medical Research office in Mwanza. Data can also be accessed through the INDEPTH Network’s repository operator, at https://www.indepth-ishare.org/index.php/catalog/78. Demographic surveillance data and data from the epidemiological sero-survey can also be found at the ‘DataFirst’ repository of the ALPHA research network, at https://www.datafirst.uct.ac.za/dataportal/index.php/catalog/903. If collaborative analyses of the data through a data-sharing agreement is desired, please contact the Centre Manager, Dr Safari Kinung’hi at.

## Acknowledgements

Thank you to the Global Fund Round 9 for providing the funds for the Sero8 survey. Thank you to the National Medical Research Coordinating Committee for providing ethical approval for the study. Finally, we extend many thanks to the study participants who generously shared personal details for the advancement of science.

